# Predicting clinical outcome with phenotypic clusters in COVID-19 pneumonia: an analysis of 12,066 hospitalized patients from the Spanish registry SEMI-COVID-19

**DOI:** 10.1101/2020.09.14.20193995

**Authors:** Manuel Rubio-Rivas, Xavier Corbella, José María Mora-Luján, Jose Loureiro Amigo, Almudena López Sampalo, Carmen Yera Bergua, Pedro Jesús Esteve Atiénzar, Luis Felipe Díez García, Ruth Gonzalez Ferrer, Susana Plaza Canteli, Antía Pérez Piñeiro, Begoña Cortés Rodríguez, Leyre Jorquer Vidal, Ignacio Pérez Catalán, Marta Leon Tellez, José Ángel Martín Oterino, María Candelaria Martín González, José Luis Serrano Carrillo de Albornoz, Eva Garcia Sardon, José Nicolás Alcalá Pedrajas, Anabel Martin-Urda Diez-Canseco, M<u>^a^</u>José Esteban Giner, Pablo Tellería Gómez, Ricardo Gómez-Huelgas, José Manuel Ramos-Rincón, for the SEMI-COVID-19 Network

## Abstract

**(1) Background:** This study aims to identify different clinical phenotypes in COVID-19 pneumonia using cluster analysis and to assess the prognostic impact among identified clusters in such patients.

**(2) Methods:** Cluster analysis including 11 phenotypic variables was performed in a large cohort of 12,066 COVID-19 patients, collected and followed-up from March 1, to July 31, 2020, from the nationwide Spanish SEMI-COVID-19 Registry.

**(3) Results:** Of the total of 12,066 patients included in the study, most were males (7,052, 58.5%) and Caucasian (10,635, 89.5%), with a mean age at diagnosis of 67 years (SD 16). The main pre-admission comorbidities were arterial hypertension (6,030, 50%), hyperlipidemia (4,741, 39.4%) and diabetes mellitus (2,309, 19.2%). The average number of days from COVID-19 symptom onset to hospital admission was 6.7 days (SD 7). The triad of fever, cough, and dyspnea was present almost uniformly in all 4 clinical phenotypes identified by clustering. Cluster C1 (8,737 patients, 72.4%) was the largest, and comprised patients with the triad alone. Cluster C2 (1,196 patients, 9.9%) also presented with ageusia and anosmia; cluster C3 (880 patients, 7.3%) also had arthromyalgia, headache, and sore throat; and cluster C4 (1,253 patients, 10.4%) also manifested with diarrhea, vomiting, and abdominal pain. Compared to each other, cluster C1 presented the highest in-hospital mortality (24.1% vs. 4.3% vs. 14.7% vs. 18.6%; p<0.001). The multivariate study identified phenotypic clusters as an independent factor for in-hospital death.

**(4) Conclusion:** The present study identified 4 phenotypic clusters in patients with COVID-19 pneumonia, which predicted the in-hospital prognosis of clinical outcomes.

## 1. Introduction

Since January 2020, the COVID-19 pneumonia pandemic has spread across the globe. As of August 13th, 2020, 20,624,830 people have been infected worldwide and 749,424 people have died. Numerous studies have highlighted the clinical characteristics of the disease [1-3]. From the beginning, different clinical forms in presentation and prognosis have been intuited; however, these clinical forms have not been defined yet. Although some factors associated with poor prognosis are known [4], it is not clear which patients may present a worse evolution during hospitalization and why.

The present study aimed to identify clinical phenotypes by cluster analysis in our large nationwide series of COVID-19 pneumonia and to create a predictive model related to a poor outcome.

## 2. Materials and methods

### 2.1. Study Design, Patient Selection, and Data Collection

A cluster analysis was performed in the large cohort of consecutive patients included in the Spanish registry SEMI-COVID-19, created by the Spanish Society of Internal Medicine (SEMI). This is a multicenter, nationwide registry with 109 hospitals registered so far. From March 1, to July 31, 126 2020, 12,066 hospitalized patients providing data of symptoms of COVID-19 upon admission were included in the Registry. All included patients were diagnosed by polymerase chain reaction (PCR) test taken from a nasopharyngeal sample, sputum or bronchoalveolar lavage.

All participating centers in the register received confirmation from the relevant Ethics 130 Committees, including Bellvitge University Hospital (PR 128/20).

### 2.2. Treatments prescribed

The treatments received were in accordance with the medical guidelines available at the time of the pandemic [5-11]. In the absence of clinical evidence of any of the treatments at the initial time of the pandemic, their use was allowed off-label.

### 2.3. Outcomes definition

The primary outcome of the study was in-hospital mortality. The secondary outcome was the requirement of mechanical ventilation or intensive care unit (ICU) admission.

### 2.4. Statistical analysis

Categorical variables were expressed as absolute numbers and percentages. Continuous variables are expressed as mean plus standard deviation (SD) in case of parametric distribution or median [IQR] in the case of non-parametric distribution. Differences among groups were assessed using the chi-square test for categorical variable and ANOVA or Kruskal-Wallis test as appropriate for continuous variables. P-values< 0.05 indicated statistical significance.

The cluster analysis was performed by ascendant hierarchical clustering on the 11 variables previously selected by using Ward’s minimum variance method with Euclidean squared distance [12]. Results are graphically depicted by a dendrogram. The number of clusters was estimated by a visual distance criterion of the dendrogram. The cluster analysis model was included in a binary logistic regression, taking the two above-mentioned outcomes as dependent variables. Mortality among the groups was represented by the Kaplan-Meier curves with their logarithmic range test.

Statistical analysis was performed by IBM SPSS Statistics for Windows, Version 26.0. Armonk, NY: IBM Corp.

## 3. Results

### 3.1. General data and symptoms

A total of 12,066 patients were included in the study. General data of the whole cohort are summarized in Table 1. Patients were mostly males (7,052, 58.5%) and Caucasian (10,635, 89.5%). The mean age at diagnosis was 67 years (SD 16). The average number of days from symptom onset to hospital admission was 6.7 days (SD 7). The main pre-admission comorbidities were arterial hypertension (6,030, 50%), hyperlipidemia (4,741, 39.4%) and diabetes mellitus (2,309, 19.2%). The mean Charlson index among patients was 1.2 (SD 1.8). The most common symptoms (Table 2) were fever 10,346 (85.7%), cough (9,142, 75.8%), dyspnea (7,205, 59.7%), arthromyalgia (3,794, 31.4%), diarrhea (2,943, 24. 4%), headache (1,402, 11.6%), sore throat (1,191, 9.9%), ageusia (992, 8.2%), vomiting (891, 7.4%), anosmia (879, 7.3%), and abdominal pain (738, 6.1%).

**Table 1.** General data between clusters

|  | All patients<br>N=12,066 | C1<br>N=8,737 | C2<br>N=1,196 | C3<br>N=880 | C4<br>N=1,253 | p-value |
| --- | --- | --- | --- | --- | --- | --- |
| Age yr, median [IQR] | 68 [56-79] | 70 [57-80] | 61 [51-71] | 64 [52-75] | 67 [53-77] | <0.001 |
| Gender, males n (%) | 7,052 (58.5) | 5,303 (60.8) | 643 (53.8) | 507 (57.6) | 599 (47.9) | <0.001 |
| Race |  |  |  |  |  |  |
| Caucasian | 10,635 (89.5) | 7,820 (90.9) | 1,023 (86.7) | 738 (84.7) | 1,054 (86) |  |
| Black | 43 (0.4) | 35 (0.4) | 3 (0.3) | 1 (0.1) | 4 (0.3) |  |
| Hispanic | 1,041 (8.8) | 643 (7.5) | 137 (11.6) | 117 (13.4) | 144 (11.7) | <0.001 |
| Asian | 59 (0.5) | 41 (0.5) | 2 (0.2) | 6 (0.7) | 10 (0.8) |  |
| Others | 100 (0.8) | 62 (9.7) | 15 (1.3) | 9 (1) | 14 (1.1) |  |
| BMI, median [IQR] | 28 [25-31] | 28 [25-31] | 28 [25-31] | 28 [25-31] | 28 [25-31] | 0.426 |
| Days from onset to admission, median [IQR] | 7 [4-9] | 6 [3-9] | 8 [6-10] | 7 [4-10] | 7 [4-9] | <0.001 |
| Smoking behaviour, n (%) |  |  |  |  |  |  |
| Never | 8,035 (69.7) | 5,761 (69.2) | 793 (68.7) | 587 (69.4) | 894 (74.3) |  |
| Current smoker | 567 (4.9) | 414 (5) | 64 (5.5) | 41 (4.8) | 48 (4) | 0.027 |
| Former smoker | 2,930 (25.4) | 2,153 (25.9) | 297 (25.7) | 218 (25.8) | 262 (21.8) |  |
| Comorbidity, n(%) |  |  |  |  |  |  |
| Arterial hypertension | 6,030 (50) | 4,571 (52.4) | 468 (39.1) | 386 (43.9) | 605 (48.4) | <0.001 |
| Diabetes mellitus | 2,309 (19.2) | 1,774 (20.4) | 177 (14.8) | 156 (17.8) | 202 (16.2) | <0.001 |
| Hyperlipidemia | 4,741 (39.4) | 3,527 (40.4) | 420 (35.1) | 325 (37) | 469 (37.5) | 0.001 |
| COPD | 786 (6.5) | 649 (7.4) | 44 (3.7) | 43 (4.9) | 50 (4) | <0.001 |
| Asthma | 869 (7.2) | 630 (7.2) | 90 (7.5) | 57 (6.5) | 92 (7.4) | 0.827 |
| OSAS | 751 (6.3) | 574 (6.6) | 57 (4.8) | 48 (5.5) | 72 (5.8) | 0.057 |
| Ischaemic cardiopathy | 931 (7.7) | 722 (8.3) | 49 (4.1) | 65 (7.4) | 95 (7.6) | <0.001 |
| Chronic heart failure | 809 (6.7) | 660 (7.6) | 41 (3.4) | 42 (4.8) | 66 (5.3) | <0.001 |
| Chronic kidney disease | 696 (5.8) | 550 (6.3) | 36 (3) | 36 (4.1) | 74 (5.9) | <0.001 |
| Chronic hepatopathy | 440 (3.7) | 330 (3.8) | 46 (3.8) | 22 (2.5) | 42 (3.4) | <0.001 |
| Active cancer | 1,196 (9.9) | 916 (10.5) | 94 (7.9) | 72 (8.2) | 114 (9.1) | 0.005 |
| Autoimmune disease | 277 (2.3) | 195 (2.2) | 33 (2.8) | 19 (2.2) | 30 (2.4) | 0.701 |
| Charlson index, median [IQR] | 1 [0-2] | 1 [0-2] | 0 [0-1] | 0 [0-1] | 0 [0-2] | <0.001 |

BMI: body mass index. COPD: chronic obstructive pulmonary disease. OSAS: obstructive sleep apnea syndrome

**Table 2.**
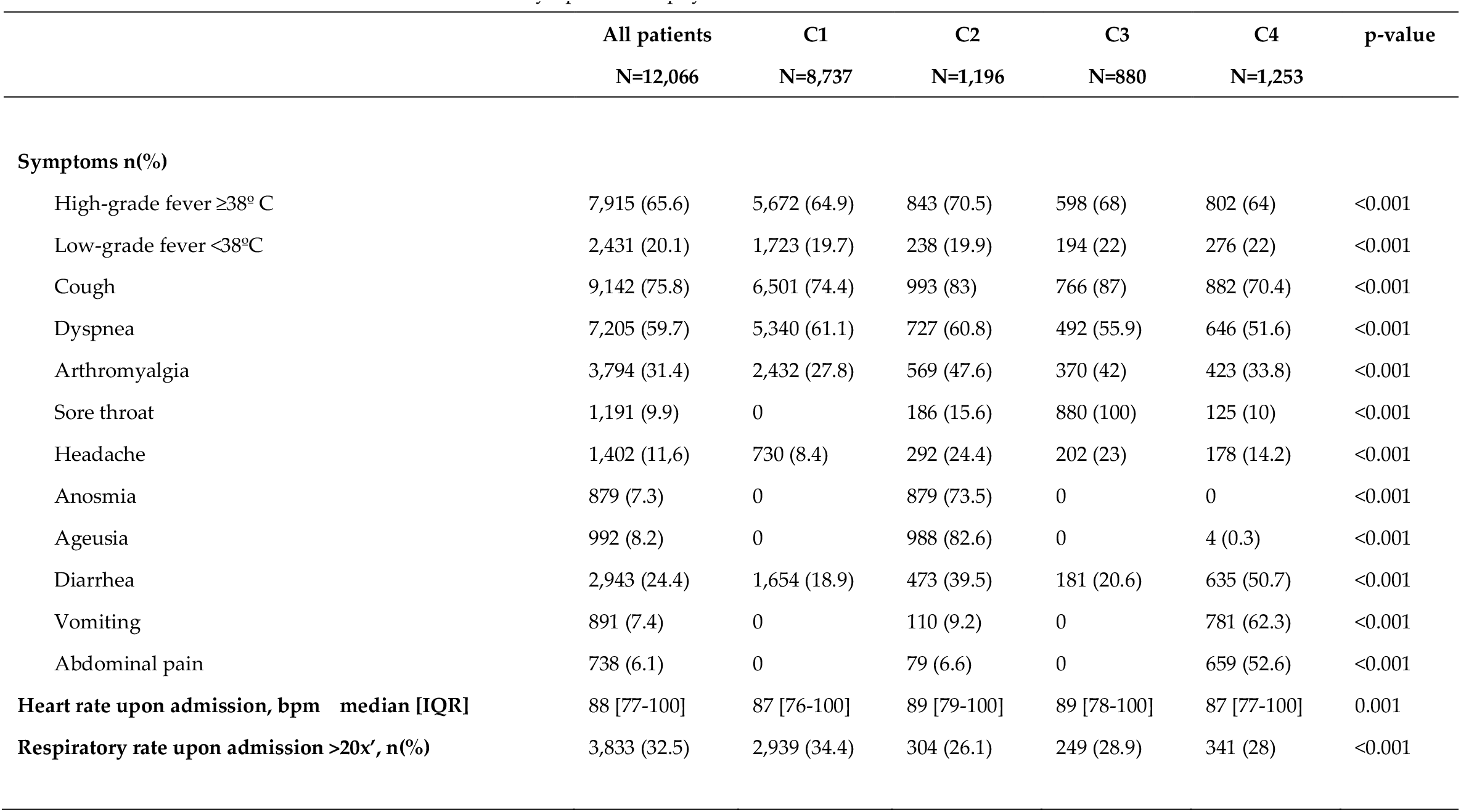
Symptoms and physical examination between clusters

### 3.2 Clustering analysis

Despite most patients presenting with fever, cough, and/or dyspnea, 4 different clusters were identified. The main characteristics of each are shown in Tables 1-5. Cluster C1 (8,737 patients, 72.4%) comprised patients with the triad of fever, cough, and dyspnea, with no other predominant symptoms. Subjects grouped in cluster C1 tended to be elderly males with a higher prevalence of comorbidities. The time between symptom onset and admission was also shorter in this subgroup of patients, in comparison with the other identified clusters. One in ten C1 patients required ICU admission and a quarter of them died, representing the highest mortality rate among the 4 clusters. Patients in the C2 cluster (1,196 patients, 9.9%) comprised patients predominantly presenting with ageusia and/or anosmia, often accompanied by fever, cough, and/or dyspnea. Subjects grouped in the C2 cluster showed the lowest percentage of ICU admission and mortality rate. Cluster C3 (880 patients, 7.3%) included patients predominantly with arthromyalgia, headache, and/or sore throat presentations, often also accompanied by fever, cough, and/or dyspnea. Up to 9.7% of C3 patients required ICU admission and 14.5% died. Finally, subjects grouped in cluster C4 (1,253 patients, 10.4%) presented predominantly with diarrhea, vomiting, and/or abdominal pain, also often accompanied by fever, cough, and/or dyspnea. Of these, 8.5% required ICU admission and 18.6% died. This mortality rate of cluster C4 was second only to the C1.

**Table 3.**
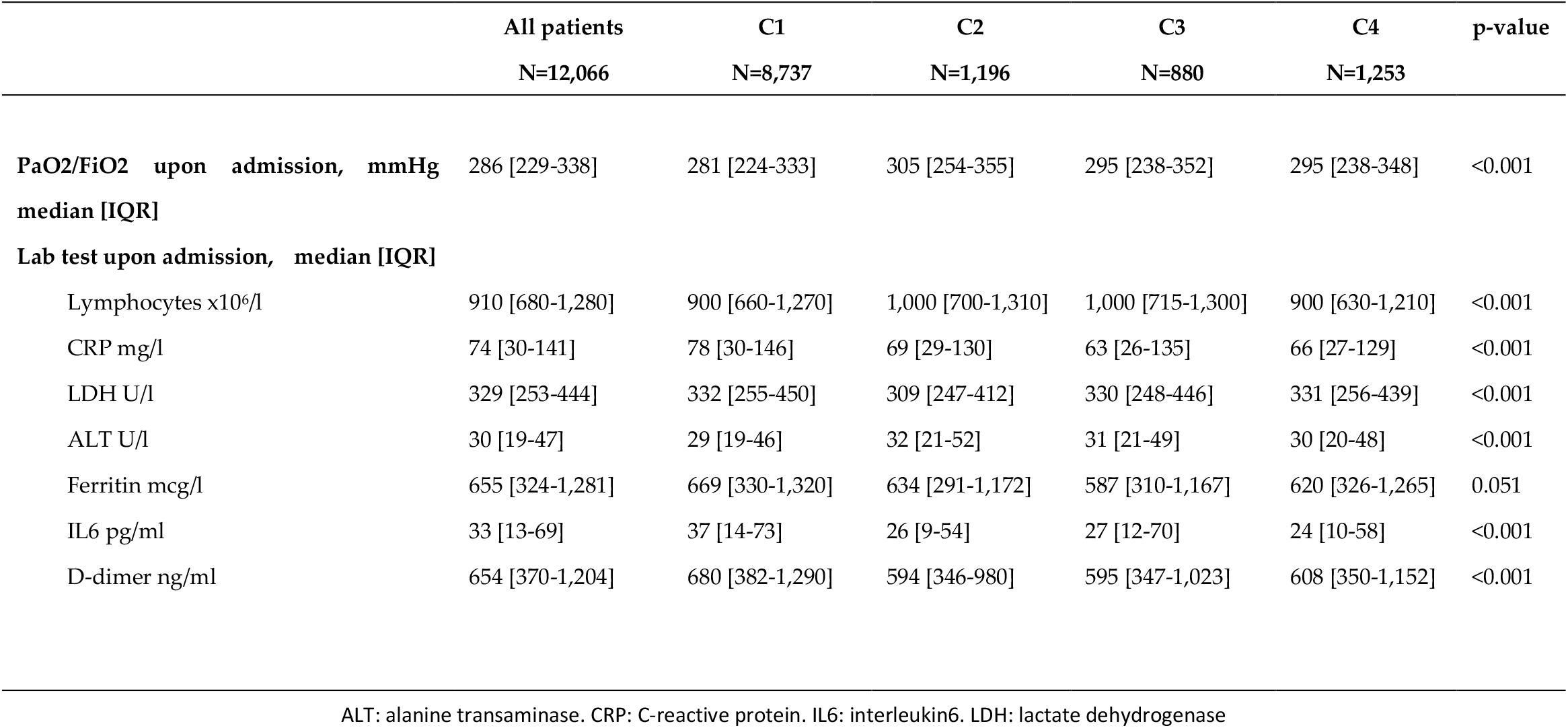
Lab tests between clusters

**Table 4.** Treatments between clusters

|  | All<br>patients<br>N=12,066 | C1<br>N=8,737 | C2<br>N=1,196 | C3<br>N=880 | C4<br>N=1,253 | p-value |
| --- | --- | --- | --- | --- | --- | --- |
| **HCQ, n (%)** | 10,665 | 7,654 (87.9) | 1,130 (94.5) | 770 (87.6) | 1,111 (88.8) | <0.001 |
| **LPV/r, n (%)** | (88.6) | 5,640 (64.8) | 783 (65.5) | 610 (69.5) | 861 (69) | 0.002 |
| **Azithromycin, n (%)** | 7,894 (65.7) | 5,407 (62.2) | 835 (69.8) | 510 (58) | 806 (64.5) | <0.001 |
| **Remdesivir, n (%)** | 7,558 (62.9) | 36 (0.4) | 10 (0.8) | 5 (0.6) | 9 (0.7) | 0.150 |
|  | 60 (0.5) |  |  |  |  |  |
| **Interferon, n (%)** | 1,496 (12.5) | 1,122 (13) | 68 (5.7) | 141 (16.1) | 165 (13.2) | <0.001 |
| **Tocilizumab, n (%)** | 1,121 (9.3) | 810 (9.3) | 110 (9.2) | 93 (10.6) | 108 (8.7) | 0.487 |
| **Corticosteroids, n (%)** | 4,343 (36.2) | 3,254 (37.5) | 399 (33.5) | 273 (31.2) | 417 (33.4) | <0.001 |
| **Heparin, n (%)** |  |  |  |  |  | <0.001 |
| Prophylactic LMWH | 7,903 (65.9) | 5,633 (65) | 817 (68.5) | 584 (66.6) | 869 (69.7) |  |
| Middle doses LMWH | 815 (6.8) | 589 (6.8) | 97 (8.1) | 49 (5.6) | 80 (6.4) |  |
| High doses LMWH | 1,305 (10.9) | 997 (11.5) | 120 (10.1) | 90 (10.3) | 98 (7.9) |  |
| **Oral anticoagulation, n (%)** |  |  |  |  |  | 0.004 |
| Oral anti-vitamin K drugs | 189 (1.6) | 156 (1.8) | 10 (0.8) | 7 (0.8) | 16 (1.3) |  |
| DOACs | 195 (1.6) | 157 (1.8) | 10 (0.8) | 10 (1.1) | 18 (1.4) |  |

DOACs: direct oral anticoagulants. HCQ: hydroxychloroquine. LPV/r: lopinavir/ritonavir. LMWH: low-molecular weight heparin

**Table 5.**
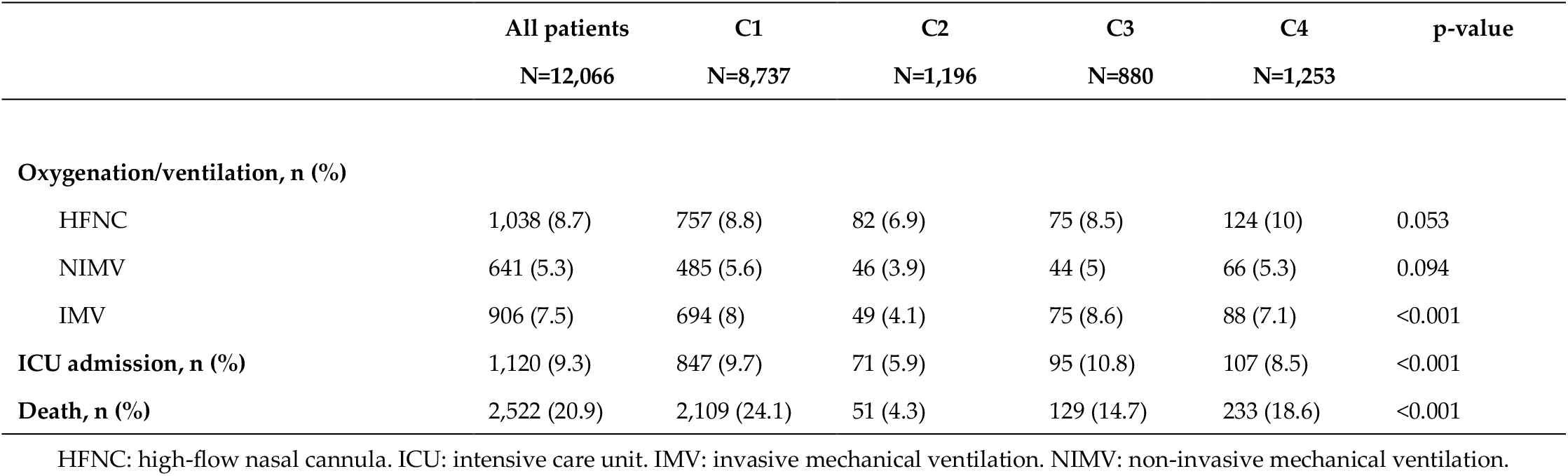
Outcomes between clusters

**Table 6.** Risk factors of in-hospital mortality.

|  | Univariate analysis<br>OR (95%CI) | p-value | Multivariate<br>analysis<br>OR (95%CI) | p-value |
| --- | --- | --- | --- | --- |
| **Age/year** | 1.09 (1.09-1.10) | <0.001 | 1.07 (1.04-1.11) | <0.001 |
| **Gender (female)** | 0.78 (0.71-0.86) | <0.001 | 0.24 (0.10-0.56) | 0.001 |
| **BMI** | 1.02 (1.01-1.04) | <0.001 | 1.09 (1.02-1.17) | 0.014 |
| **Comorbidity** |  |  |  |  |
| Arterial hypertension | 3.07 (2.79-3.38) | <0.001 | NS |  |
| Diabetes mellitus | 2.07 (1.87-2.29) | <0.001 | NS |  |
| Hyperlipidemia | 1.80 (1.64-1.96) | <0.001 | NS |  |
| COPD | 2.82 (2.43-3.27) | <0.001 | NS |  |
| Ischaemic cardiopathy | 2.67 (2.32-3.07) | <0.001 | NS |  |
| Chronic heart failure | 3.74 (3.23-4.32) | <0.001 | NS |  |
| Chronic kidney disease | 3.18 (2.72-3.72) | <0.001 | NS |  |
| Chronic hepatopathy | 1.57 (1.27-1.94) | <0.001 | NS |  |
| Active cancer | 2.23 (1.96-2.53) | <0.001 | NS |  |
| **Charlson index** | 1.37 (1.34-1.41) | <0.001 | 1.52 (1.30-1.78) | <0.001 |
| **Heart rate upon admission** | 1.00 (0.99-1.00) | 0.278 |  |  |
| **Respiratory rate upon admission >20x'** | 4.48 (4.08-4.92) | <0.001 | 2.84 (1.33-6.05) | 0.007 |
| **PaO2/FiO2 upon admission** | 0.99 (0.99-0.99) | <0.001 | 0.99 (0.98-1.00) | 0.001 |
| **Lab test upon admission** |  |  |  |  |
| Lymphocytes x10<sup>6</sup>/l | 1.00 (1.00-1.00) | 0.768 |  |  |
| CRP mg/l | 1.01 (1.01-1.01) | <0.001 | 0.99 (0.99-1.00) | 0.034 |
| LDH U/l | 1.00 (1.00-1.00) | <0.001 | 1.00 (1.00-1.00) | 0.032 |
| ALT U/l | 1.00 (0.99-1.00) | 0.792 |  |  |
| Ferritin mcg/l | 1.00 (1.00-1.00) | <0.001 | 1.00 (1.00-1.00) | 0.001 |
| IL6 pg/ml | 1.00 (1.00-1.00) | <0.001 | 1.00 (1.00-1.00) | 0.020 |
| D-dimer ng/ml | 1.00 (1.00-1.00) | <0.001 | 1.00 (1.00-1.00) | 0.054 |
| **Treatments during admission** |  |  |  |  |
| Remdesivir | 1.16 (0.64-2.12) | 0.623 |  |  |
| Tocilizumab | 1.24 (1.07-1.43) | 0.004 | NS |  |
| Corticosteroids | 2.06 (1.89-2.26) | <0.001 | NS |  |
| **Clusters** |  |  |  |  |
| C1 | 1 ref. |  | 1 ref. |  |
| C2 | 0.14 (0.11-0.19) | <0.001 | 0.91 (0.30-2.75) | 0.865 |
| C3 | 0.54 (0.45-0.66) | <0.001 | 0.18 (0.04-0.96) | 0.044 |
| C4 | 0.72 (0.62-0.84) | <0.001 | 2.85 (0.88-9.22) | 0.082 |

BMI: body mass index. COPD: chronic obstructive pulmonary disease.

Analytical results among clusters showed that PaO2/FiO2 at entry was a median 286 mmHg [229-338], being highest in the C2 cluster (281 mmHg vs. 305 vs. 295 vs. 295; p<0.001). Cluster C1 showed the highest values of C-reactive protein (CRP) (78 mg/l vs. 69 vs. 63 vs. 66; p<0.001), lactate dehydrogenate (LDH) (332 U/l vs. 309 vs. 330 vs. 331; p<0.001), ferritin (669 mcg/l vs. 634 vs. 587 vs. 620; p=0.051), interleukin-6 (IL-6) (37 pg/ml vs. 26 vs. 27 vs. 24; p<0.001), and D-dimer (680 ng/ml vs. 594 vs. 595 vs. 608; p<0.001).

### 3.3 Treatments and outcomes

The treatments received are shown in Table 4. As antiviral treatment, patients were treated with hydroxychloroquine (HCQ) (10,665, 88.6%), Lopinavir/ritonavir (LPV/r) (7,894, 65.7%), azithromycin (7,558, 62.9%) and remdesivir (60, 0.5%). As immunomodulatory treatments, they received corticosteroids (4,343, 36.2%), interferon (1,496, 12.5%) and tocilizumab (1,121, 9.3%). As anticoagulant treatment, patients received oral anticoagulation (384, 3.18%) or low-molecular-weight heparin (LMWH) at prophylactic doses (7,903, 65.9%), intermediate doses (815, 6.8%) or full doses (1,305, 10.9%).

Of the total 12,066, 1,038 (8.7%) patients required high-flow nasal cannula (HFNC), 641 (5.3%) non-invasive mechanical ventilation (NIMV), and 906 (7.5%) invasive mechanical ventilation (IMV). Admissions to the ICU numbered 1,120 patients (9.3%). Overall, the mortality rate was 20.9% (2,522 patients). The outcomes are shown in Table 5.

### 3.4 Predictive model for mortality

A predictive study of uni- and multivariate logistic regression using in-hospital death as a dependent variable was performed. The predictors of mortality in the multivariate study were as follows: age [OR 1.07 (1.04-1.11)], gender (female) [OR 0.24 (0.10-0.56)], BMI [OR 1.09 (1.02-1. 17)], Charlson index [OR 1.52 (1.30-1.78)], respiratory rate upon admission >20 bpm [OR 2.84 (1.33-6.05)], PaO2/FiO2 upon admission [OR 0.99 (0.98-1.00)], CRP [OR 0.99 (0.99-1.00)], LDH [OR 1.00 (1.00-1.00)], ferritin [OR 1.00 (1.00-1.00)], IL-6 [OR 1.00 (1.00-1.00)], and the phenotypic cluster. The C1 cluster was chosen as a reference. Clusters C2 [OR 0.91 (0.30-2.31)] and C3 [OR 0.18 (0.04-0.96)] had a better prognosis in the multivariate study. The C4 cluster was also observed to have a poor prognosis [OR 2.85 (0.88-9.22)].

**Figure 1.**
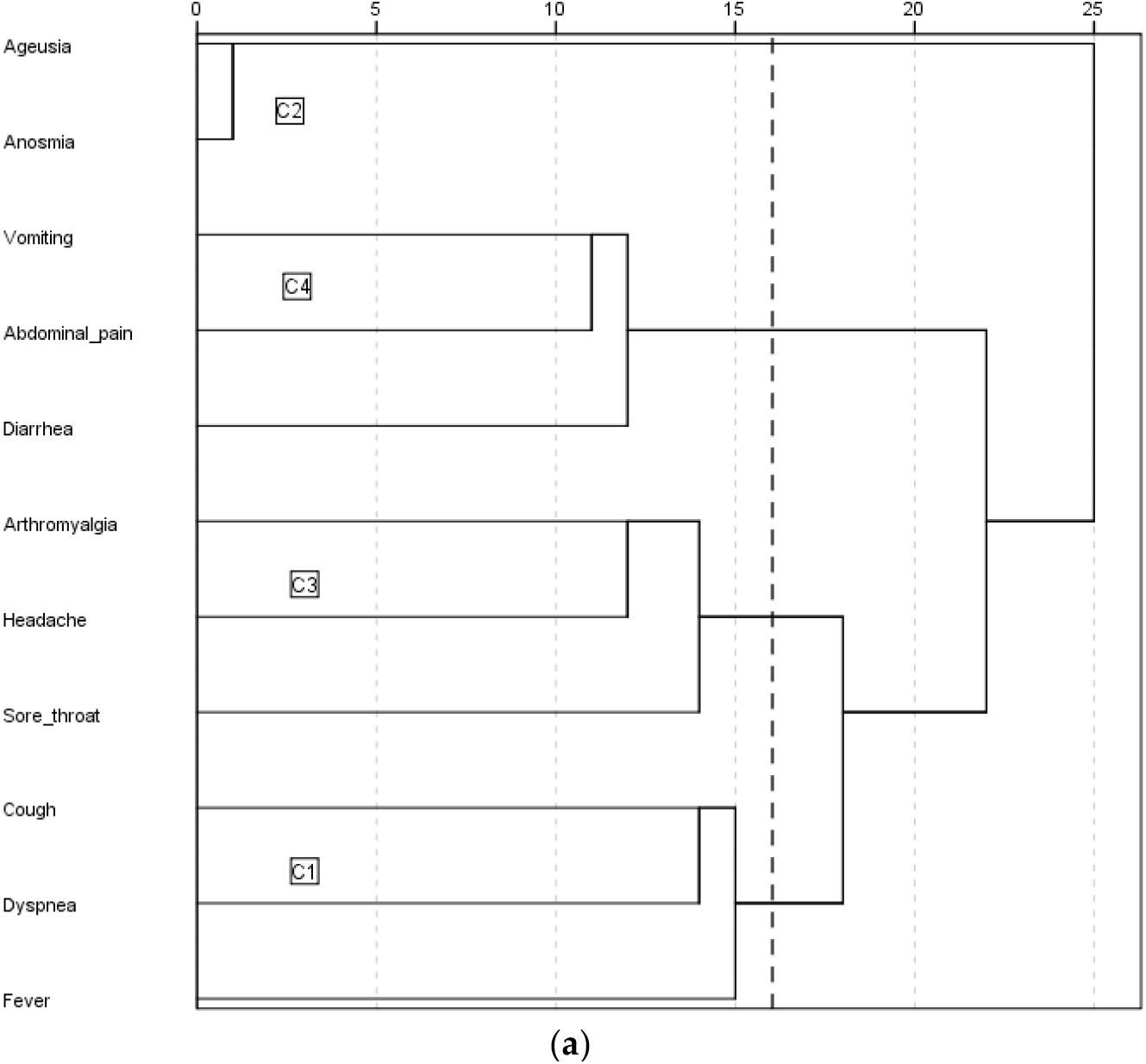
Dendrogram.

**Figure 2.**
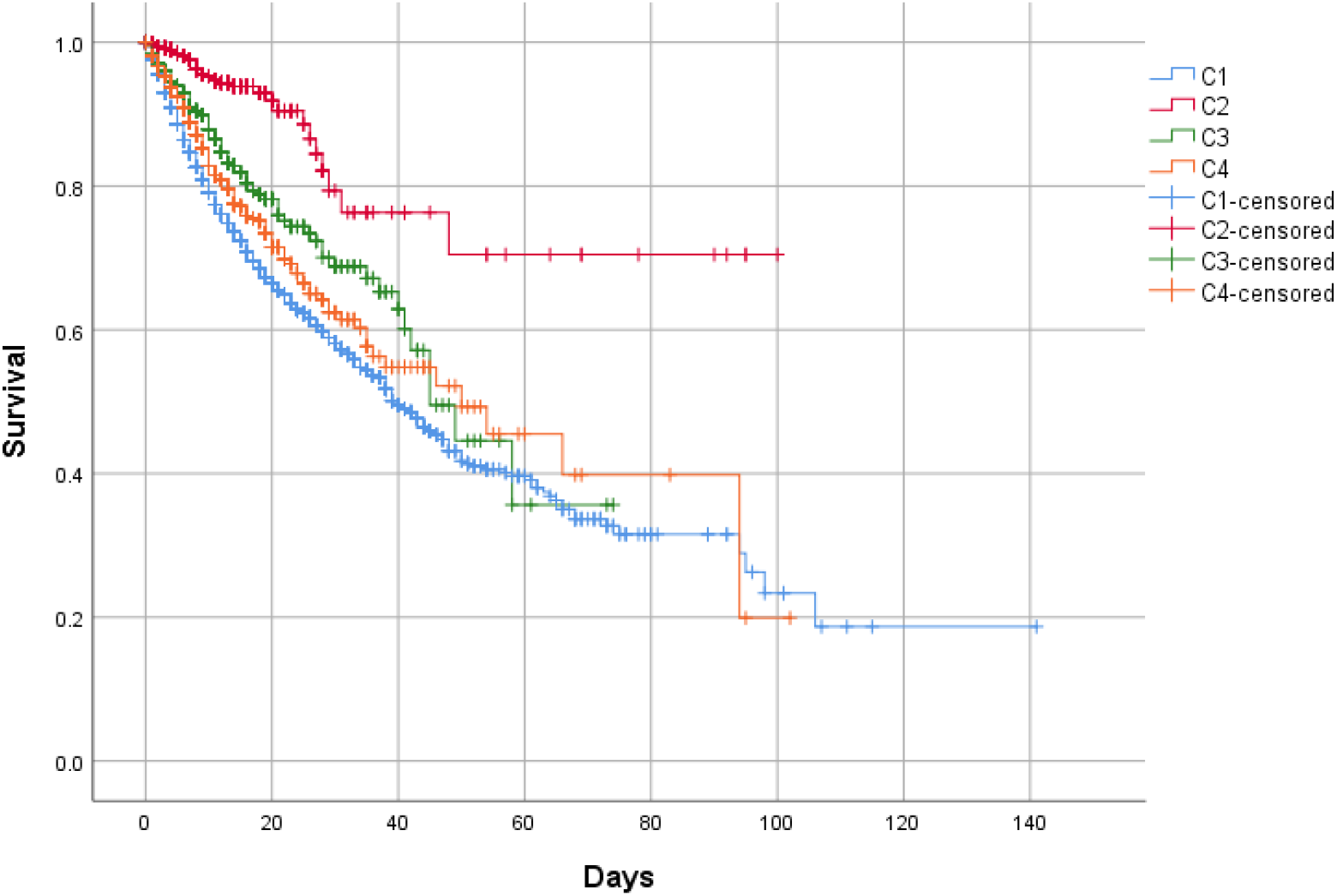
In-hospital mortality between clusters. Kaplan-Meier. Log-rank test p<0.001

## 4. Discussion

The present investigation shows data from the first study of phenotypic clusters in COVID-19 pneumonia. The source of data was the Spanish registry SEMI-COVID-19, whose characteristics have recently been published [13]. Our analysis showed the existence of 4 clusters with differentiated clinical peculiarities and different prognoses.

The general characteristics of age, gender, and comorbidities found in our study are consistent with those already described in the literature. Likewise, the treatments administered are in accordance with the study period covered by the record.

The triad of fever, cough, and dyspnea was present almost uniformly in all patients with COVID-19 pneumonia grouped in the 4 phenotypes. However, other particular symptoms may help clinicians to differentiate them. Cluster C1 does not usually present symptoms in addition to the triad of fever, cough, and dyspnea. Subjects grouped in the C2 cluster usually present with ageusia and/or anosmia in addition to the triad. Cluster C3 is characterized by the presence of concomitant arthromyalgia, headache, and/or sore throat. Finally, the C4 cluster also manifests with digestive symptoms such as diarrhea, vomiting, and/or abdominal pain.

In terms of prognosis, the C1 cluster showed the highest mortality rate (24.1%) in this large Spanish nation-wide series. It was followed by C4 (18.6%), C3 (14.7%), and finally C2 (4.3%). The crude survival study identified the C2 cluster as a cluster of good prognosis. The multivariate regression study showed a non-significative trend to better prognosis. Also identified the C3 cluster as another good prognostic subgroup, in addition to C2. In contrast, the C1 and C4 clusters were identified as the poorest prognosis clusters.

The risk factors recognized so far for poor prognosis have been repeated in several studies. The mainly reported risk factors are advanced age, male gender, higher BMI, and some analytical parameters such as PaO2/FiO2, lymphocyte count, CRP, LDH, ferritin, IL-6, and D-dimer. Certain comorbidities such as diabetes mellitus, arterial hypertension, or hyperlipidemia have also been suggested as poor prognostic factors but not identified to date.

Interestingly, the study presented here identifies the cluster phenotype as a new prognostic factor. Since clusters share common characteristics, sometimes it can be difficult to recognize which cluster a patient belongs to. However, in other many occasions, the clinical profile may be sufficiently evident to recognize the cluster, helping physicians to make clinical decisions based on prognostic information of the identified cluster.

To date, there are no published, peer-reviewed phenotypic cluster studies in the medical literature on COVID-19. A study of clusters in out-of-hospital population can be found in the medRxiv repository [14]. It is based on an app in which patients enter their symptoms. With these data and some other clinical data provided by the patient, a risk of respiratory support (defined as the need for oxygen therapy or mechanical ventilation) is deduced. It is therefore a predictor of hospitalization, we could say. We have some doubts as to whether the source of the data can be considered reliable since the data is not introduced by a doctor but by the patient himself. On the other hand, the fact that it is based on an app may represent a bias against the elderly population not accustomed to electronic devices. They identify 6 phenotypic clusters, with some similarity and overlap with the clusters presented in our study. It is an interesting tool, specially designed for general practitioners.

As for the generalization of our results, it should be noted that the data come from a developed European western country with a mostly Caucasian population and little representation of other ethnicities. Furthermore, it should also be taken into account that Spain has a universal-coverage public healthcare system, not comparable with some other developed and developing countries. On the other hand, proportionally speaking, Spain has one of the largest elderly populations in the world and, as is well known, age has been described as a fundamental factor in the poor prognosis of COVID-19 pneumonia [4]. These characteristics could influence the outcomes shown.

In order to speak properly, the definition of a true phenotype requires a consistent natural history, similar clinical and physiological characteristics, underlying pathobiology with identifiable biomarkers and genetics, and predictable response to general and specific therapies [15]. Accordingly, it would be necessary to study each of the present clinical clusters genetically and to verify that each cluster has a differentiated genetic background. In the literature, some studies attempted to phenotype patients with COVID-19 as a function of the immune response, and others suggested phenotyping as a function of pathophysiology [16,17]. It would be interesting to combine all methods of phenotyping.

We believe that the identification of the present clusters may be of great help to clinicians in order to identify those cases with a better or worse prognosis, and thus direct more individualized therapeutic strategies. In this regard, we also believe that identification of phenotypes can serve as a guide for clinical trials, not evaluating new treatments in general, since not all subgroups of COVID-19 patients may benefit from the same therapeutic strategies. On the other hand, drugs previously discarded, but with a rational pathophysiological basis to be tested, should be reanalyzed to clarify their real efficacy, taking into account the different clinical spectrum of COVID-19 patients.

The main strength of this study is the identification of different phenotypic clusters in COVID-19 pneumonia from a very large sample of more than 12,000 patients from more than 100 hospitals. Among limitations, data were obtained from a retrospective register of a sole country, which means that some specific data could be missing or collected with some grade of heterogeneity.

## 5. Conclusions

In conclusion, the present study identified 4 phenotypic clusters that predicted in-hospital prognosis of clinical outcome in a large nationwide series of patients with COVID-19 pneumonia. Clusters associated with bad in-hospital prognosis were C1, in which subjects presented with the isolated triad of fever, cough, and dyspnea, and C4 also manifested with diarrhea, vomiting, and/or abdominal pain. In contrast, subjects grouped in the C2 cluster (manifested also with ageusia and/or anosmia) showed the best prognosis, together with cluster C3 (adding arthromyalgia, headache, and/or sore throat), which was second only to C2 showing a good outcome.

## Data Availability

The datasets generated during and/or analyzed during the current study are available from the corresponding author on reasonable request

## Acknowledgments

We gratefully acknowledge all the investigators who participate in the SEMI-COVID-19 Registry. We also thank the SEMI-COVID-19 Registry Coordinating Center, S&H Medical Science Service, for their quality control data, logistic and administrative support. The authors declare that there are no conflicts of interest.

## Author Contributions

For research articles with several authors, a short paragraph specifying their individual contributions must be provided. The following statements should be used "Conceptualization, M.R.-R., X.C and J.M. M.-L.; methodology, M.R.; software, M.R.; validation, M.R.-R., X.C.; formal analysis, M.R.-R.; investigation, M.R.-R., X.C.; resources, R.G.-H., J.M.R.-R.; data curation, M.R.-R., X.C, J.M. M.-L., J.L.A., A.L.S., C.Y.B., V.G.G., L.F.D.G., R.G.F., S.P.C., S.F.C., B. C.R., L.J.V., I.P.C., M.L.T., J.A.M.O., M.C.M.G., J.L.S.C., E.G.S., J.N.A.P., A.M.-U.D.-C., M.J.E.G., P.T.G., R.G.-H., J.M.R.-R.; writing—original draft preparation, M.R.-R., X.C.; writing—review and editing, M.R.-R., X.C.; visualization M.R.-R., X.C.; supervision, M.R.-R., X.C, R.G.-H., J.M.R.-R.; project administration, J.M.R.-R. All authors have read and agreed to the published version of the manuscript.", please turn to the CRediT taxonomy for the term explanation. Authorship must be limited to those who have contributed substantially to the work reported.

## Funding

This research received no external funding

## Conflicts of Interest

The authors declare no conflict of interest.

## Appendix. List of the SEMI-COVID-19 Network members

Coordinator of the SEMI-COVID-19 Registry: José Manuel Casas Rojo.

SEMI-COVID-19 Scientific Committee Members: José Manuel Casas Rojo, José Manuel Ramos Rincón, Carlos Lumbreras Bermejo, Jesús Millán Núñez-Cortés, Juan Miguel Antón Santos, Ricardo Gómez Huelgas.

SEMI-COVID-19 Registry Coordinating Center: S & H Medical Science Service.

### Members of the SEMI-COVID-19 Group

#### H. U. 12 de Octubre. Madrid

Paloma Agudo de Blas, Coral Arévalo Cañas, Blanca Ayuso, José Bascuñana Morejón, Samara Campos Escudero, María Carnevali Frías, Santiago Cossio Tejido, Borja de Miguel Campo, Carmen Díaz Pedroche, Raquel Diaz Simon, Ana García Reyne, Lucia Jorge Huerta, Antonio Lalueza Blanco, Jaime Laureiro Gonzalo, Carlos Lumbreras Bermejo, Guillermo Maestro de la Calle, Barbara Otero Perpiña, Diana Paredes Ruiz, Marcos Sánchez Fernández, Javier Tejada Montes.

#### H. U. Gregorio Marañón. Madrid

Laura Abarca Casas, Alvaro Alejandre de Oña, Rubén Alonso Beato, Leyre Alonso Gonzalo, Jaime Alonso Muñoz, Crhistian Mario Amodeo Oblitas, Cristina Ausín García, Marta Bacete Cebrián, Jesús Baltasar Corral, Maria Barrientos Guerrero, Alejandro Bendala Estrada, María Calderón Moreno, Paula Carrascosa Fernández, Raquel Carrillo, Sabela Castañeda Pérez, Eva Cervilla Muñoz, Agustín Diego Chacón Moreno, Maria Carmen Cuenca Carvajal, Sergio de Santos, Andrés Enríquez Gómez, Eduardo Fernández Carracedo, María Mercedes Ferreiro-Mazón Jenaro, Francisco Galeano Valle, Alejandra Garcia, Irene Garcia Fernandez-Bravo, María Eugenia García Leoni, Maria Gomez Antunez, Candela González San Narciso, Anthony Alexander Gurjian, Lorena Jiménez Ibáñez, Cristina Lavilla Olleros, Cristina Llamazares Mendo, Sara Luis García, Víctor Mato Jimeno, Clara Millán Nohales, Jesús Millán Núñez-Cortés, Sergio Moragón Ledesma, Antonio Muiño Miguez, Cecilia Muñoz Delgado, Lucía Ordieres Ortega, Susana Pardo Sánchez, Alejandro Parra Virto, María Teresa Pérez Sanz, Blanca Pinilla Llorente, Sandra Piqueras Ruiz, Guillermo Soria Fernández-Llamazares, María Toledano Macías, Neera Toledo Samaniego, Ana Torres do Rego, Maria Victoria Villalba Garcia, Gracia Villarreal, María Zurita Etayo.

#### Hospital Universitari de Bellvitge. L’Hospitalet de Llobregat

Xavier Corbella, Narcís Homs, Abelardo Montero, Jose María Mora-Luján, Manuel Rubio-Rivas.

#### H. U. La Paz-Cantoblanco-Carlos III. Madrid

Jorge Álvarez Troncoso, Francisco Arnalich Fernández, Francisco Blanco Quintana, Carmen Busca Arenzana, Sergio Carrasco Molina, Aranzazu Castellano Candalija, Germán Daroca Bengoa, Alejandro de Gea Grela, Alicia de Lorenzo Hernández, Alejandro Díez Vidal, Carmen Fernández Capitán, Maria Francisca García Iglesias, Borja González Muñoz, Carmen Rosario Herrero Gil, Juan María Herrero Martínez, Víctor Hontañón, Maria Jesús Jaras Hernández, Carlos Lahoz, Cristina Marcelo Calvo, Juan Carlos Martín Gutiérrez, Monica Martinez Prieto, Elena Martínez Robles, Araceli Menéndez Saldaña, Alberto Moreno Fernández, Jose Maria Mostaza Prieto, Ana Noblejas Mozo, Carlos Manuel Oñoro López, Esmeralda Palmier Peláez, Marina Palomar Pampyn, Maria Angustias Quesada Simón, Juan Carlos Ramos Ramos, Luis Ramos Ruperto, Aquilino Sánchez Purificación, Teresa Sancho Bueso, Raquel Sorriguieta Torre, Clara Itziar Soto Abanedes, Yeray Untoria Tabares, Marta Varas Mayoral, Julia Vásquez Manau.

#### C. H. U. de Albacete. Albacete

Jose Luis Beato Pérez, Maria Lourdes Sáez Méndez.

#### Complejo Asistencia! de Segovia. Segovia

Eva María Ferreira Pasos, Daniel Monge Monge, Alba Varela García.

#### H. U. Puerta de Hierro. Majadahonda

María Álvarez Bello, Ane Andrés Eisenhofer, Ana Arias Milla, Isolina Baños Pérez, Javier Bilbao Garay, Silvia Blanco Alonso, Jorge Calderón Parra, Alejandro Callejas Díaz, José María Camino Salvador, M<u>^a^</u> Cruz Carreño Hernández, Valentín Cuervas-Mons Martínez, Sara de la Fuente Moral, Miguel del Pino Jimenez, Alberto Díaz de Santiago, Itziar Diego Yagüe, Ignacio Donate Velasco, Ana María Duca, Pedro Durán del Campo, Gabriela Escudero López, Esther Expósito Palomo, Ana Fernández Cruz, Esther Fiz Benito, Andrea Fraile López, Amy Galán Gómez, Sonia García Prieto, Claudia García Rodríguez-Maimón, Miguel Ángel García Viejo, Javier Gómez Irusta, Edith Vanessa Gutiérrez Abreu, Isabel Gutiérrez Martín, Ángela Gutiérrez Rojas, Andrea Gutiérrez Villanueva, Jesús Herráiz Jiménez, Pedro Laguna del Estal, M<u>^a^</u> Carmen Máinez Sáiz, Cristina Martín Martín, María Martínez Urbistondo, Fernando Martínez Vera, Susana Mellor Pita, Patricia Mills Sánchez, Esther Montero Hernández, Alberto Mora Vargas, Cristina Moreno López, Alfonso Ángel-Moreno Maroto, Victor Moreno-Torres Concha, Ignacio Morrás De La Torre, Elena Múñez Rubio, Ana Muñoz Gómez, Rosa Muñoz de Benito, Alejandro Muñoz Serrano, Jose María Palau Fayós, Ilduara Pintos Pascual, Antonio Ramos Martínez, Isabel Redondo Cánovas del Castillo, Alberto Roldán Montaud, Lucía Romero Imaz, Yolanda Romero Pizarro, Mónica Sánchez Santiuste, David Sánchez Órtiz, Enrique Sánchez Chica, Patricia Serrano de la Fuente, Pablo Tutor de Ureta, Ángela Valencia Alijo, Mercedes Valentín-Pastrana Aguilar, Juan Antonio Vargas Núñez, Jose Manuel Vázquez Comendador, Gema Vázquez Contreras, Carmen Vizoso Gálvez.

#### H. Miguel Servet. Zaragoza

Gonzalo Acebes Repiso, Uxua Asín Samper, María Aranzazu Caudevilla Martínez, José Miguel García Bruñén, Rosa García Fenoll, Jesús Javier González Igual, Laura Letona Giménez, Mónica Llorente Barrio, Luis Sáez Comet.

#### H. U. La Princesa. Madrid

María Aguilera García, Ester Alonso Monge, Jesús Álvarez Rodríguez, Claudia Alvarez Varela, Miquel Berniz Gòdia, Marta Briega Molina, Marta Bustamante Vega, Jose Curbelo, Alicia de las Heras Moreno, Ignacio Descalzo Godoy, Alexia Constanza Espiño Alvarez, Ignacio Fernández Martín-Caro, Alejandra Franquet López-Mosteiro, Gonzalo Galvez Marquez, María J. García Blanco, Yaiza García del Álamo Hernández, Clara García-Rayo Encina, Noemí Gilabert González, Carolina Guillamo Rodríguez, Nicolás Labrador San Martín, Manuel Molina Báez, Carmen Muñoz Delgado, Pedro Parra Caballero, Javier Pérez Serrano, Laura Rabes Rodríguez, Pablo Rodríguez Cortés, Carlos Rodriguez Franco, Emilia Roy-Vallejo, Monica Rueda Vega, Aresio Sancha Lloret, Beatriz Sánchez Moreno, Marta Sanz Alba, Jorge Serrano Ballester, Alba Somovilla, Carmen Suarez Fernández, Macarena Vargas Tirado, Almudena Villa Marti.

#### H. U. de A Coruña. A Coruña

Alicia Alonso Álvarez, Olaya Alonso Juarros, Ariadna Arévalo López, Carmen Casariego Castiñeira, Ana Cerezales Calviño, Marta Contreras Sánchez, Ramón Fernández Varela, Santiago J. Freire Castro, Ana Padín Trigo, Rafael Prieto Jarel, Fátima Raad Varea, Laura Ramos Alonso, Francisco Javier Sanmartín Pensado, David Vieito Porto.

#### H. Clínico San Carlos. Madrid

Inés Armenteros Yeguas, Javier Azaña Gómez, Julia Barrado Cuchillo, Irene Burruezo López, Noemí Cabello Clotet, Alberto E. Calvo Elías, Elpidio Calvo Manuel, Carmen María Cano de Luque, Cynthia Chocron Benbunan, Laura Dans Vilan, Ester Emilia Dubon Peralta, Vicente Estrada Pérez, Santiago Fernandez-Castelao, Marcos Oliver Fragiel Saavedra, José Luis García Klepzig, Maria del Rosario Iguarán Bermúdez, Esther Jaén Ferrer, Rubén Ángel Martín Sánchez, Manuel Méndez Bailón, Maria José Nuñez Orantos, Carolina Olmos Mata, Eva Orviz García, David Oteo Mata, Cristina Outon González, Juncal Perez-Somarriba, Pablo Pérez Mateos, Maria Esther Ramos Muñoz, Xabier Rivas Regaira, Iñigo Sagastagoitia Fornie, Alejandro Salinas Botrán, Miguel Suárez Robles, Maddalena Elena Urbano, Miguel Villar Martínez.

#### H. Infanta Sofía. S. S. de los Reyes

Rafael del Castillo Cantero, Rebeca Fuerte Martínez, Arturo Muñoz Blanco, José Francisco Pascual Pareja, Isabel Perales Fraile, Isabel Rábago Lorite, Llanos Soler Rangel, Inés Suárez García, Jose Luis Valle López.

#### Hospital Royo Villanova. Zaragoza

Nicolás Alcalá Rivera, Anxela Crestelo Vieitez, Esther del Corral, Jesús Díez Manglano, Isabel Fiteni Mera, Maria del Mar Garcia Andreu, Martin Gerico Aseguinolaza, Claudia Josa Laorden, Raul Martinez Murgui, Marta Teresa Matía Sanz.

#### H. Moisès Broggi. Sant Toan Despí

Judit Aranda Lobo, Jose Loureiro Amigo, Isabel Oriol Bermúdez, Melani Pestaña Fernández, Nicolas Rhyman, Nuria Vázquez Piqueras.

#### Hospital Universitario Dr. Peset. Valencia

Juan Alberto Aguilera Ayllón, Arturo Artero, María del Mar Carmona Martín, María José Fabiá Valls, Maria de Mar Fernández Garcés, Ana Belén Gómez Belda, Ian López Cruz, Manuel Madrazo López, Elisabet Mateo Sanchis, Jaume Micó Gandia, Laura Piles Roger, Adela Maria Pina Belmonte, Alba Viana García.

#### Hospital Clínico de Santiago. Santiago de Compostela

Maria del Carmen Beceiro Abad, Maria Aurora Freire Romero, Sonia Molinos Castro, Emilio Manuel Paez Guillan, María Pazo Nuñez, Paula Maria Pesqueira Fontan.

#### H. Nuestra Señora del Prado. Talavera de la Reina

Sonia Casallo Blanco, Jeffrey Oskar Magallanes Gamboa.

#### H. U. Ramón y Cajal. Madrid

Luis Fernando Abrego Vaca, Ana Andréu Arnanz, Octavio Arce García, Marta Bajo González, Pablo Borque Sanz, Alberto Cozar Llisto, Sonia de Pedro Baena, Beatriz Del Hoyo Cuenda, María Alejandra Gamboa Osorio, Isabel García Sánchez, Andrés González García, Oscar Alberto López Cisneros, Miguel Martínez Lacalzada, Borja Merino Ortiz, Jimena Rey-García, Elisa Riera González, Cristina Sánchez Díaz, Grisell Starita Fajardo, Cecilia Suárez Carantoña, Adrian Viteri Noel, Svetlana Zhilina Zhilina.

#### H. U. Infanta Cristina. Parla

Juan Miguel Antón Santos, Ana Belén Barbero Barrera, Coralia Bueno Muiño, Ruth Calderón Hernaiz, Irene Casado Lopez, José Manuel Casas Rojo, Andrés Cortés Troncoso, Mayte de Guzmán García-Monge, Francesco Deodati, Gonzalo García Casasola Sánchez, Elena Garcia Guijarro, Davide Luordo, María Mateos González, Jose A Melero Bermejo, Lorea Roteta García, Elena Sierra Gonzalo, Javier Villanueva Martínez.

#### H. de Cabueñes. Gijón

Ana María Alvarez Suárez, Carlos Delgado Vergés, Rosa Fernandez-Madera Martínez, Eva Fonseca Aizpuru, Alejandro Gómez Carrasco, Cristina Helguera Amezua, Juan Francisco López Caleya, María del Mar Martínez López, Aleida Martínez Zapico, Carmen Olabuenaga Iscar, María Luisa Taboada Martínez, Lara María Tamargo Chamorro.

#### Hospital de Urduliz Alfredo Espinosa. Urdúliz

María Aparicio López, Asier Aranguren Arostegui, Paula Arriola Martínez, Gorka Arroita Gonzalez, M<u>^a^</u> Soledad Azcona Losada, Miriam García Gómez, Eduardo Garcia Lopez, Amalur Iza Jiménez, Alazne Lartategi Iraurgi, Esther Martinez Becerro, Itziar Oriñuela González, Isabel María Portales Fernández, Pablo Ramirez Sánchez, Beatriz Ruiz Estévez, Cristian Vidal Núñez.

#### H. Virgen de la Salud. Toledo

Ana Maria Alguacil Muñoz, Marta Blanco Fernández, Veronica Cano, Ricardo Crespo Moreno, Fernando Cuadra Garcia-Tenorio, Blanca Díaz-Tendero Nájera, Raquel Estévez González, María Paz García Butenegro, Alberto Gato Díez, Verónica Gómez Caverzaschi, Piedad María Gómez Pedraza, Julio González Moraleja, Raúl Hidalgo Carvajal, Patricia Jiménez Aranda, Raquel Labra González, Áxel Legua Caparachini, Pilar Lopez Castañeyra, Agustín Lozano Ancin, Jose Domingo Martin Garda, Cristina Morata Romero, María Jesús Moya Saiz, Helena Moza Moríñigo, Gemma Muñiz Nicolás, Enriqueta Muñoz Platon, Filomena Oliveri, Elena Ortiz Ortiz, Raúl Perea Rafael, Pilar Redondo Galán, María Antonia Sepulveda Berrocal, Vicente Serrano Romero de Ávila, Pilar Toledano Sierra, Yamilex Urbano Aranda, Jesús Vázquez Clemente, Carmen Yera Bergua.

#### Hospital Regional Universitario de Málaga. Málaga

M<u>^a^</u> Mar Ayala Gutiérrez, Rosa Bernal López, José Bueno Fonseca, Verónica Andrea Buonaiuto, Luis Francisco Caballero Martínez, Lidia Cobos Palacios, Clara Costo Muriel, Francis de Windt, Ana Teresa Fernandez-Truchaud Christophel, Paula García Ocaña, Ricardo Gómez Huelgas, Javier Gorospe García, Maria Dolores López Carmona, Pablo López Quirantes, Almudena López Sampalo, Elizabeth Lorenzo Hernández, Juan José Mancebo Sevilla, Jesica Martin Carmona, Luis Miguel Pérez-Belmonte, Araceli Pineda Cantero, Michele Ricci, Jaime Sanz Cánovas

#### H. Santa Marina. Bilbao

Maria Areses Manrique, Ainara Coduras Erdozain, Ane Elbire Labirua-Iturburu Ruiz.

#### Hospital HLA Moncloa. Madrid

Teresa Garcia Delange, Isabel Jimenez Martinez, Carmen Martinez Cilleros, Nuria Parra Arribas.

#### H. del Henares. Coslada

Jesús Ballano Rodríguez-Solís, Luis Cabeza Osorio, María del Pilar Fidalgo Montero, M<u>^a^</u> Isabel Fuentes Soriano, Erika Esperanza Lozano Rincon, Ana Martín Hermida, Jesus Martinez Carrilero, Jose Angel Pestaña Santiago, Manuel Sánchez Robledo, Patricia Sanz Rojas, Nahum Jacobo Torres Yebes, Vanessa Vento.

#### H. U. Torrevieja. Torrevieja

Julio César Blázquez Encinar, Joaquín Fernández López-Cuervo.

#### H. U. La Fe. Valencia

Dafne Cabañero, María Calabuig Ballester, Pascual Císcar Fernández, Ricardo Gil Sánchez, Marta Jiménez Escrig, Cristina Marín Amela, Laura Parra Gómez, Carlos Puig Navarro, José Antonio Todolí Parra.

#### H. San Pedro. Logroño

Diana Alegre González, Irene Ariño Pérez de Zabalza, Sergio Arnedo Hernández, Jorge Collado Sáenz, Beatriz Dendariena, Marta Gómez del Mazo, Iratxe Martínez de Narvajas Urra, Sara Martínez Hernández, Estela Menendez Fernández, Jose Luís Peña Somovilla, Elisa Rabadán Pejenaute.

#### Hospital Universitario Ntra Sra Candelaria. Santa Cruz de Tenerife

Lucy Abella, Andrea Afonso Díaz, Selena Gala Aguilera Garcia, Marta Bethencourt Feria, Eduardo Mauricio Calderón Ledezma, Sara Castaño Perez, Guillermo Castro Gainett, José Manuel del Arco Delgado, Joaquín Delgado Casamayor, Diego Garcia Silvera, Alba Gómez Hidalgo, Marcelino Hayek Peraza, Carolina Hernández Carballo, Rubén Hernández Luis, Francisco Javier Herrera Herrera, Maria del Mar Lopez Gamez, Julia Marfil Daza, María José Monedero Prieto, María Blanca Monereo Muñoz, María de la Luz Padilla Salazar, Daniel Rodríguez Díaz, Alicia Tejera, Laura Torres Hernández.

#### H. U. San Juan de Alicante. San Juan de Alicante

David Balaz, David Bonet Tur, Pedro Jesús Esteve Atiénzar, Carles García Cervera, David Francisco García Núñez, Vicente Giner Galvañ, Angie Gómez Uranga, Javier Guzmán Martínez, Isidro Hernández Isasi, Lourdes Lajara Villar, Juan Manuel Núñez Cruz, Sergio Palacios Fernández, Juan Jorge Peris García, Andrea Riaño Pérez, José Miguel Seguí Ripoll, Philip Wikman-Jorgensen.

#### H. U. San Agustin. Avilés

Andrea Álvarez García, Víctor Arenas García, Alba Barragán Mateos, Demelsa Blanco Suárez, María Caño Rubia, Jaime Casal Álvarez, David Castrodá Copa, José Ferreiro Celeiro, Natalia García Arenas, Raquel García Noriega, Joaquin Llorente García, Irene Maderuelo Riesco, Paula Martinez Garcia, Maria Jose Menendez Calderon, Diego Eduardo Olivo Aguilar, Marta Nataya Solís Marquínez, Luis Trapiella Martínez, Andrés Astur Treceño García, Juan Valdés Bécares.

#### H. de Mataró. Mataró

Raquel Aranega González, Ramon Boixeda, Carlos Lopera Mármol, Marta Parra Navarro, Ainhoa Rex Guzmán, Aleix Serrallonga Fustier.

#### H. U. Son Llatzer. Palma de Mallorca

Andrés de la Peña Fernández, Almudena Hernández Milián.

#### H. Juan Ramón Jiménez. Huelva

Francisco Javier Bejarano Luque, Francisco Javier Carrasco-Sánchez, Mercedes de Sousa Baena, Jaime Díaz Leal, Aurora Espinar Rubio, Maria Franco Huertas, Juan Antonio García Bravo, Andrés Gonzalez Macías, Encarnación Gutiérrez Jiménez, Alicia Hidalgo Jiménez, Constantino Lozano Quintero, Carmen Mancilla Reguera, Francisco Javier Martínez Marcos, Francisco Muñoz Beamud, Maria Perez Aguilera, Alícia Perez Jiménez, Virginia Rodríguez Castaño, Alvaro Sánchez de Alcazar del Río, Leire Toscano Ruiz.

#### H. U. Reina Sofía. Córdoba

Antonio Pablo Arenas de Larriva, Pilar Calero Espinal, Javier Delgado Lista, María Jesús Gómez Vázquez, Jose Jiménez Torres, Laura Martín Piedra, Javier Pascual Vinagre, María Elena Revelles Vílchez, Juan Luis Romero Cabrera, José David Torres Peña.

#### Hospital Infanta Margarita. Cabra

María Esther Guisado Espartero, Lorena Montero Rivas, Maria de la Sierra Navas Alcántara, Raimundo Tirado-Miranda.

#### H. U. Virgen de las Nieves. Granada

Pablo Conde Baena, Joaquin Escobar Sevilla, Laura Gallo Padilla, Patricia Gómez Ronquillo, Pablo González Bustos, María Navío Botías, Jessica Ramírez Taboada, Mar Rivero Rodríguez.

#### Hospital Costa del Sol. Marbella

Victoria Augustín Bandera, María Dolores Martín Escalante.

Complejo Asistencial Universitario de León. León

Rosario Maria García Diez, Manuel Martin Regidor, Angel Luis Martínez Gonzalez, Alberto Muela Molinero, Raquel Rodríguez Díez, Beatriz Vicente Montes.

#### Hospital Marina Baixa. Villajoyosa

Javier Ena, Jose Enrique Gómez Segado.

#### C. H. U. de Ferrol. Ferrol

Hortensia Alvarez Diaz, Tamara Dalama Lopez, Estefania Martul Pego, Carmen Mella Pérez, Ana Pazos Ferro, Sabela Sánchez Trigo, Dolores Suarez Sambade, Maria Trigas Ferrin, Maria del Carmen Vázquez Friol, Laura Vilariño Maneiro.

#### Hospital Torrecárdenas. Almería

Luis Felipe Díez García, Iris El Attar Acedo, Bárbara Hernandez Sierra, Carmen Mar Sánchez Cano.

#### Hospital Clinic Barcelona. Barcelona

Júlia Calvo Jiménez, Aina Capdevila Reniu, Irene Carbonell De Boulle, Emmanuel Coloma Bazán, Joaquim Fernández Sola, Cristina Gabara Xancó, Joan Ribot Grabalosa, Olga Rodríguez Núñez.

#### Hospital del Tajo. Aranjuez

Ruth Gonzalez Ferrer, Raquel Monsalvo Arroyo.

#### Hospital Insular de Gran Canaria. Las Palmas G. C

Marina Aroza Espinar, Jorge Orihuela Martín, Carlos Jorge Ripper, Selena Santana Jiménez.

#### H. U. Severo Ochoa. Leganés

Yolanda Casillas Viera, Lucía Cayuela Rodríguez, Carmen de Juan Alvarez, Gema Flox Benitez, Laura García Escudero, Juan Martin Torres, Patricia Moreira Escriche, Susana Plaza Canteli, M Carmen Romero Pérez.

#### Hospital Alto Guadalquivir. Andújar

Begoña Cortés Rodríguez.

#### Hospital Valle del Nalón. Riaño (Langreo)

Sara Fuente Cosío, César Manuel Gallo Álvaro, Julia Lobo García, Antía Pérez Piñeiro.

#### H. Francesc de Borja. Gandia

Alba Camarena Molina, Simona Cioaia, Anna Ferrer Santolalia, José María Frutos Pérez, Eva Gil Tomás, Leyre Jorquer Vidal, Marina Llopis Sanchis, M Ángeles Martínez Pascual, Alvaro Navarro Batet, Mari Amparo Perea Ribis, Ricardo Peris Sanchez, José Manuel Querol Ribelles, Silvia Rodriguez Mercadal, Ana Ventura Esteve.

#### H. U. del Vinalopó. Elche

Francisco Amorós Martínez, Erika Ascuña Vásquez, Jose Carlos Escribano Stablé, Adriana Hernández Belmonte, Ana Maestre Peiró, Raquel Martínez Goñi, M. Carmen Pacheco Castellanos, Bernardino Soldan Belda, David Vicente Navarro.

#### H. G. U. de Castellón. Castellón de la Plana

Jorge Andrés Soler, Marián Bennasar Remolar, Alejandro Cardenal Álvarez, Daniela Díaz Carlotti, María José Esteve Gimeno, Sergio Fabra Juana, Paula García López, María Teresa Guinot Soler, Daniela Palomo de la Sota, Guillem Pascual Castellanos, Ignacio Pérez Catalán, Celia Roig Martí, Paula Rubert Monzó, Javier Ruiz Padilla, Nuria Tornador Gaya, Jorge Usó Blasco.

#### C. H. U. de Badajoz. Badajoz

Rafael Aragon Lara, Inmaculada Cimadevilla Fernandez, Juan Carlos Cira García, Gema Maria García García, Julia Gonzalez Granados, Beatriz Guerrero Sánchez, Francisco Javier Monreal Periáñez, Maria Josefa Pascual Perez.

#### H. Santa Bárbara. Soria

Marta Leon Tellez.

#### C.A. U. de Salamanca. Salamanca

Gloria María Alonso Claudio, Víctor Barreales Rodríguez, Cristina Carbonell Muñoz, Adela Carpio Pérez, María Victoria Coral Orbes, Daniel Encinas Sánchez, Sandra Inés Revuelta, Miguel Marcos Martín, José Ignacio Martín González, José Ángel Martín Oterino, Leticia Moralejo Alonso, Sonia Peña Balbuena, María Luisa Pérez García, Ana Ramon Prados, Beatriz Rodríguez-Alonso, Ángela Romero Alegría, Maria Sanchez Ledesma, Rosa Juana Tejera Pérez.

#### H. U. Quironsalud Madrid. Pozuelo de Alarcón (Madrid)

Pablo Guisado Vasco, Ana Roda Santacruz, Ana Valverde Muñoz.

#### H. U. de Canarias. Santa Cruz de Tenerife

Julio Cesar Alvisa Negrin, José Fernando Armas González, Lourdes González Navarrete, Iballa Jiménez, María Candelaria Martín González, Esther Martín Ponce, Miguel Nicolas Navarrete Lorite, Paula Ortega Toledo, Onán Pérez Hernández, Alina Pérez Ramírez.

#### H. U. del Sureste. Arganda del Rey

Jon Cabrejas Ugartondo, Ana Belén Mancebo Plaza, Arturo Noguerado Asensio, Bethania Pérez Alves, Natalia Vicente López.

#### H. de Poniente. Almería

Juan Antonio Montes Romero, Encarna Sánchez Martín, Jose Luis Serrano Carrillo de Albornoz, Manuel Jesus Soriano Pérez.

#### H. Parc Tauli. Sabadell

Francisco Epelde, Isabel Torrente

#### H. San Pedro de Alcántara. Cáceres

Angela Agea Garcia, Javier Galán González, Luis Gámez Salazar, Eva Garcia Sardon, Antonio González Nieto, Itziar Montero Díaz, Selene Núñez Gaspar, Alvaro Santaella Gomez.

#### H. de Pozoblanco. Pozoblanco

José Nicolás Alcalá Pedrajas, Antonia Márquez García, Inés Vargas.

#### H. Virgen de los Lirios. Alcoy (Alicante)

M<u>^a^</u> José Esteban Giner.

#### Hospital Doctor José Molina Orosa. Arrecife (Lanzarote)

Virginia Herrero García, Berta Román Bernal.

#### Hospital de Palamós. Palamós

Maricruz Almendros Rivas, Miquel Hortos Alsina, Anabel Martin-Urda Diez-Canseco.

#### Hospital Clínico Universitario de Valladolid. Valladolid

Xjoylin Teresita Egües Torres, Sara Gutiérrez González, Cristina Novoa Fernández, Pablo Tellería Gómez.

#### H. U. Puerta del Mar. Cádiz

José Antonio Girón González, Susana Fabiola Pascual Perez, Cristina Rodríguez Fernández-Viagas, Maria José Soto Cardenas.

#### Hospital de Montilla. Montilla

Ana Cristina Delgado Zamorano, Beatriz Gómez Marín, Adrián Montaño Martínez, Jose Luis Zambrana García.

#### H. Infanta Elena. Huelva

María Gloria Rojano Rivero.

#### H. Virgen del Mar. Madrid

Thamar Capel Astrua, Paola Tatiana Garcia Giraldo, Maria Jesus Gonzalez Juarez, Victoria Marquez Fernandez, Ada Viviana Romero Echevarry.

#### Hospital do Salnes. Vilagarcía de Arousa

Vanesa Alende Castro, Ana María Baz Lomba, Ruth Brea Aparicio, Marta Fernandez Morales, Jesus Manuel Fernandez Villar, Maria Teresa Lopez Monteagudo, Cristina Pérez García, Lorena María Rodríguez Ferreira, Diana Sande Llovo, Maria Begoña Valle Feijoo.

## Notes

### Competing Interest Statement

The authors have declared no competing interest.

### Funding Statement

The authors declare that they have no conflict of interest.

### Author Declarations

All participating centers in the register received confirmation from the relevant Ethics Committees, including Bellvitge University Hospital (PR 128/20).

